# Use of electronic health record data to identify hospital-associated *Clostridioides difficile* infections: a validation study

**DOI:** 10.1101/2024.01.10.24301118

**Authors:** Michael J. Ray, Kathleen L. Lacanilao, Maela Robyne Lazaro, Luke C. Strnad, Jon P. Furuno, Kelly Royster, Jessina C. McGregor

## Abstract

**Background:** Clinical research focused on the burden and impact of *Clostridioides difficile* infection (CDI) often relies upon accurate identification of cases using existing health record data. Use of diagnosis codes alone can lead to misclassification of cases. Our goal was to develop and validate a multi-component algorithm to identify hospital-associated CDI (HA-CDI) cases using electronic health record (EHR) data.

**Methods:** We performed a validation study using a random sample of adult inpatients at a large academic hospital setting in Portland, Oregon from January 2018 to March 2020. We excluded patients with CDI on admission and those with short lengths of stay (< 4 days). We tested a multi-component algorithm to identify HA-CDI; case patients were required to have received an inpatient course of metronidazole, oral vancomycin, or fidaxomicin and have at least one of the following: a positive *C. difficile* laboratory test or the International Classification of Diseases, Tenth Revision (ICD-10) code for non-recurrent CDI. For a random sample of 80 algorithm-identified HA-CDI cases and 80 non-cases, we performed manual EHR review to identify gold standard of HA-CDI diagnosis. We then calculated overall percent accuracy, sensitivity, specificity, and positive and negative predictive value for the algorithm overall and for the individual components.

**Results:** Our case definition algorithm identified HA-CDI cases with 94% accuracy (95% Confidence Interval (CI): 88% to 97%). We achieved 100% sensitivity (94% to 100%), 89% specificity (81% to 95%), 88% positive predictive value (78% to 94%), and 100% negative predictive value (95% to 100%). Requiring a positive *C. difficile* test as our gold standard further improved diagnostic performance (97% accuracy [93% to 99%], 93% PPV [85% to 98%]).

**Conclusions:** Our algorithm accurately detected true HA-CDI cases from EHR data in our patient population. A multi-component algorithm performs better than any isolated component. Requiring a positive laboratory test for *C. difficile* strengthens diagnostic performance even further. Accurate detection could have important implications for CDI tracking and research.

## Background

*Clostridioides difficile* is responsible for nearly a third of all antibiotic-associated cases of infectious diarrhea worldwide, and healthcare-associated *C. difficile* infection (HA-CDI) represents about two-thirds of all CDI cases in the United States (1, 2). The early 2010s saw a greater than 50 percent increase in CDI incidence, which can be attributed, at least in part, to more sensitive but less specific molecular testing methods compared to previously used toxin-based assays (3-5). Over diagnosis of CDI (i.e., treating *C. difficile* colonized patients without active infection) leads to unnecessary antibiotic prescribing, often resulting in extra costs, adverse side effects, possible emergence of antibiotic resistance, and increased risk of recurrent CDI (6-8). Nevertheless, single-component case finding methods (e.g. laboratory test or diagnosis code only) are often employed for their utility. For example, since 2009, the National Healthcare Safety Network (NHSN) has utilized the LabID event, which allows for reporting of positive *C. difficile* lab events “without clinical evaluation” (9). While this is used as a proxy for CDI burden and as an inter-facility comparison tool, it likely overestimates the number of CDI cases, and has been shown to have a low negative predictive value (NPV) of around 55 percent (10). Using laboratory testing as the sole diagnostic criterion has also been shown to have low specificity (11, 12).

Our study objective was to develop and validate a CDI case definition algorithm that accurately detects CDI cases (i.e., maximizes diagnostic performance) using electronic health record (EHR) data including antibiotic treatment, laboratory test, and International Classification of Diseases, Clinical Modification (ICD-10-CM) information. We hypothesized that our case definition algorithm would detect patients with CDI more accurately than any single case definition component.

## Methods

### Study Design and Data Source

We obtained complete laboratory, pharmacy, and diagnostic code information on all Oregon Health & Science University (OHSU) inpatient hospital encounters between January 2018 and March 2020 from our institution’s research data repository, which includes longitudinal administrative, demographic, diagnosis, laboratory, and pharmacy data at the patient level that has been validated and used in previous epidemiologic studies (16-18). OHSU is a 576-bed academic, quaternary-care facility in Portland, Oregon. We established our study cohort by excluding patients under the age of 18, those with known recurrent or community-acquired CDI, and those with hospital stays of less than four calendar days, as these individuals are not eligible to be diagnosed with HA-CDI.

*Hospital-associated CDI*

Our goal was to accurately identify incident, non-recurrent HA-CDI, which we identified using a combination of medication (metronidazole, oral vancomycin, fidaxomicin), diagnosis code (ICD-10-CM (A04.72: Enterocolitis due to *Clostridioides difficile*, not specified as recurrent), and laboratory testing data (Figure 1). We considered incident CDI cases to be HA-CDI if the onset date, defined as the date of first anti-*C. difficile* antibiotic administration or *C. difficile* positive stool specimen, whichever occurred first, fell on hospital day 4 or later. This is consistent with US Centers for Disease Control and Prevention HA-CDI definition for HA-CDI (9). We considered cases non-recurrent if no prior CDI events were identified at our institution in the 8 weeks before the index CDI diagnosis applying the same diagnostic criteria.

**Figure 1.**
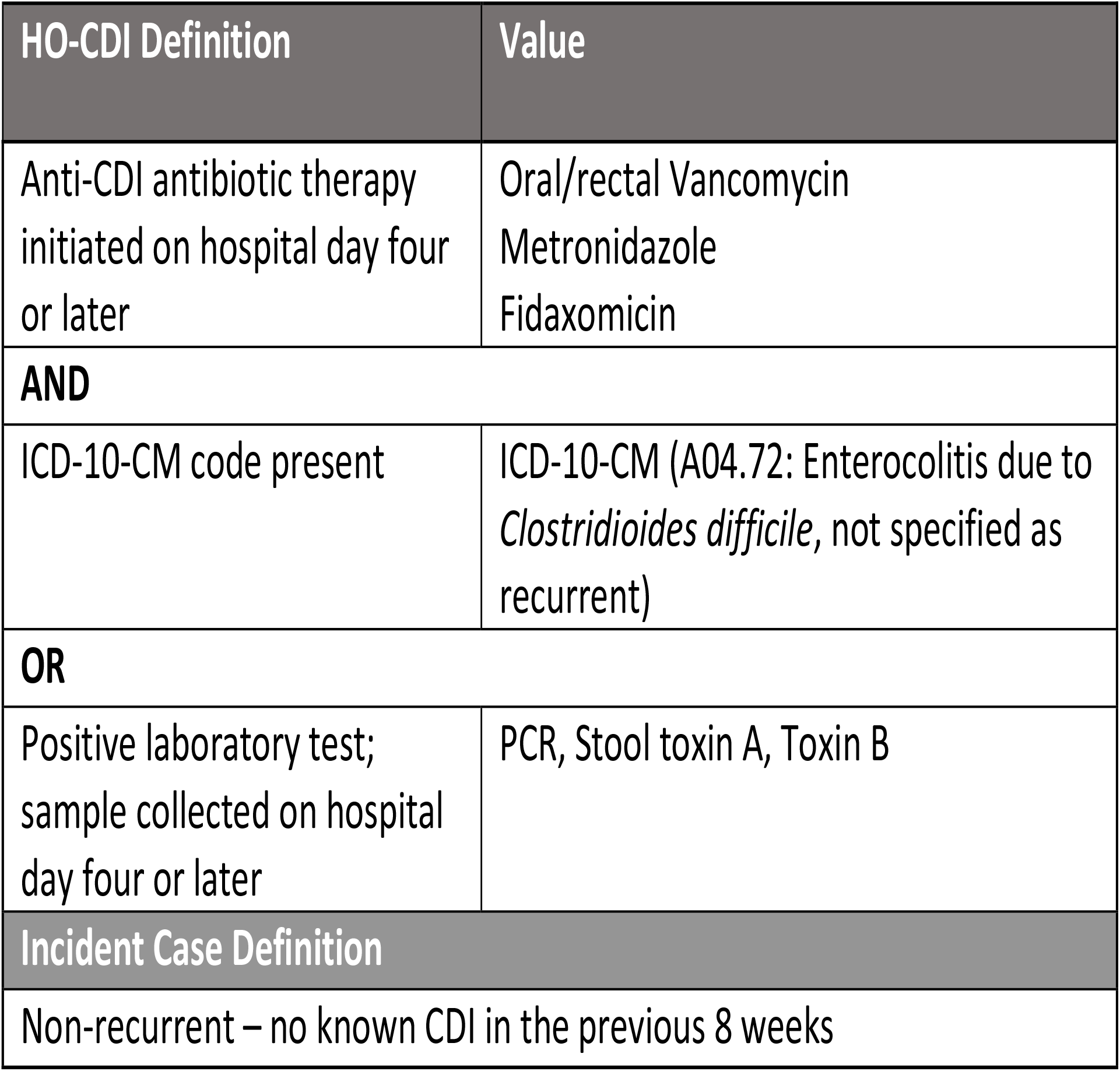
Definition for incident hospital-onset CDI cases

### Diagnostic Performance of Case Definition Algorithm

To evaluate the diagnostic performance of our case definition algorithm, we took a random sample of 80 HA-CDI cases and 80 non-CDI cases as classified by our algorithm. We determined through an *a priori* power calculation using methods outlined by Pepe and Longton (19, 20), assuming a true positive rate and false positive rate of 90 percent and 5 percent respectively and alternative true and false positive rates of 75 percent and 25 percent respectively, that this sample size would be sufficient to achieve 94% power. We (MJR, KLL, MRL, KR) then manually reviewed each encounter medical record (Epic EHR system) as our gold standard to determine if the patient was a “true” HA-CDI case. Each reviewer examined clinical notes for documentation of *C. difficile*-related diarrhea (loose, liquid, or unformed stools) and related symptoms (fever, nausea, vomiting, abdominal pain). We examined patient output assessment to determine the number of liquid stools prior to *C. difficile* testing, if applicable. We also captured information on diarrhea-inducing medications (e.g., laxatives, colchine, antineoplastic agents) both prior to and during admission. We determined if there was a documented reason for diarrhea symptoms other than *C. difficile* (e.g. tube feeding, NPO (nothing by mouth), gastrointestinal surgery) or underlying conditions that cause diarrhea (diabetic gastroparesis, ulcerative colitis, irritable bowel syndrome, Crohn’s disease). Finally, we searched the EHR for any documentation of history of CDI or CDI prophylaxis. To be considered a true HA-CDI patient and establish our gold standard, there must have been documentation of, on hospital day 4 or later, at least three loose/liquid/unformed stools with no alternative explanation documented for diarrhea symptoms, initiation of CDI-specific antibiotic treatment, and a positive laboratory test for *C. difficile* (if present). According to our review process, patients could be ruled as one of the following: HA-CDI (new CDI diagnosis on hospital day 4 or later, no known CDI in previous 8 weeks), CA-CDI (CDI diagnosis on hospital day 3 or earlier, no known CDI in previous 8 weeks), recurrent CDI (Known CDI in previous 8 weeks or documentation of recurrent CDI in chart), colonized (Positive test for *C. difficile* with the clinical decision not to treat with antibiotics), or no evidence of CDI. If any uncertainty arose as to the patient’s CDI status, we flagged the record for second review by an infectious disease specialist (LCS, KR). We utilized REDCap tools to collect and manage study data (21, 22).

### Data Analysis

Once we determined each study patient’s chart-confirmed CDI status, we calculated our case definition algorithm’s sensitivity, specificity, positive predictive value (PPV), negative predictive value (NPV), and overall percent accuracy (i.e. percent of cases and non-cases correctly identified). We constructed 95% confidence intervals (CI) using methods described by Pepe (19). We also examined the diagnostic performance of individual components of the case definition algorithm for comparison (e.g., laboratory test only, diagnosis code only, oral vancomycin only). We performed all analysis using SAS (version 9.4, SAS Corporation, Cary, NC) except for the *a priori* power calculation, for which we used Stata (version 16, StataCorp., College Station, TX).

## Results

Of the 103,275 inpatient encounters during our study period, 50,394 (49%) met our inclusion criteria (Figure 2). Included patients had a mean age of 56.7 year (standard deviation, ± 19.0 years), 53% were female, and where 87% white race. The median length of stay was 6 days (interquartile range, 5 to 9 days). Ten percent of our included patients received a CDI treatment antibiotic (metronidazole, oral vancomycin, or fidaxomicin), 710 (1.4%) received oral vancomycin or fidaxomicin, 396 (0.8%) tested positive for *C. difficile*, and 487 (1%) had an ICD-10 code for non-recurrent CDI. Of the 190 HA-CDI cases identified in our dataset, 157 (83%) had all three components of our case definition algorithm. Of the 80 algorithm-identified HA-CDI cases that we sampled for review, 66 (83%) had all three criteria, while 9 (11%) had a test and no code, and 5 (6%) had a code and no test.

**Figure 2.**
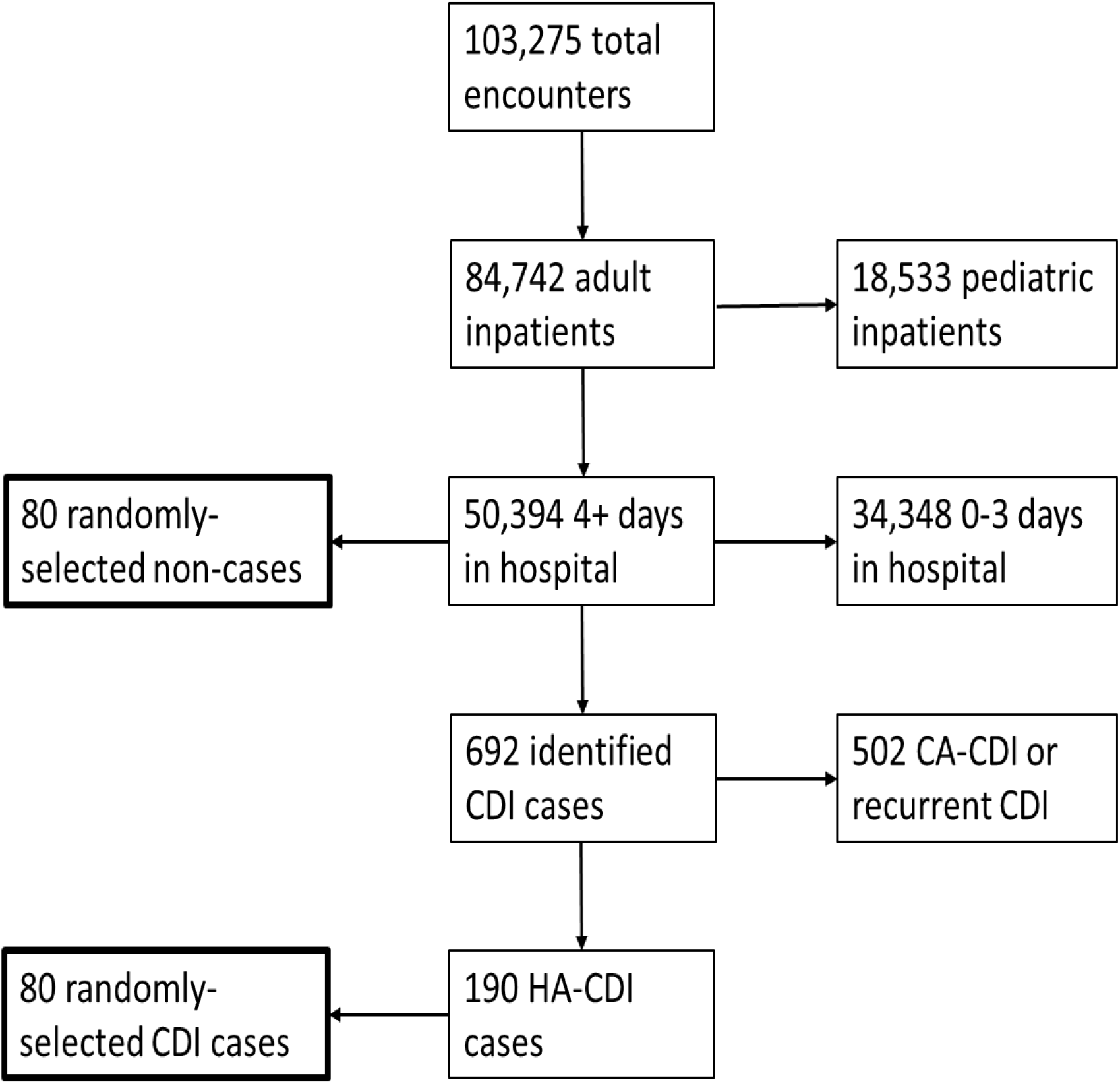
Inclusion/exclusion criteria and sampling scheme for our validation study

Our case definition algorithm identified HA-CDI cases with 94% accuracy (95% CI: 88% to 97%). We achieved 100% sensitivity (94% to 100%), 89% specificity (81% to 95%), 88% PPV (78% to 94%), and 100% NPV (95% to 100%). The performance of the individual components of our CDI algorithm is summarized in Table 1. Requiring a positive test to be considered gold-standard CDI positive (as opposed to an optional positive test if an ICD-10 code for CDI was included) improved diagnostic performance across all measures by avoiding 5 false positives, compared to the original algorithm. This improved specificity to 94% (87% to 98%), PPV to 93% (84% to 98%), and overall accuracy to 97% (93% to 99%). Requiring an ICD-10 code for CDI avoided 5 false negatives, but also resulted in 5 false positives. This reduces sensitivity and NPV, improves specificity and PPV, with the overall accuracy remaining the same. CDI specific treatment plus an ICD-10 code yielded an overall accuracy of 93% (88% to 97%), though with reduced sensitivity and NPV. Using only *C. difficile* testing yielded 15 false positives and an overall accuracy of 91% (85% to 95%), and using oral vancomycin prescribing only yielded 19 false positives, and an overall accuracy of 88% (82% to 93%).

**Table 1.** Diagnostic performance of our CDI algorithm and comparison of individual algorithm components.

| Diagnostic criteria | Diagnostic performance measure, % (95% CI) |  |
| --- | --- | --- |
| **<u>Study algorithm:</u>**<br>*ICD-10-CM code A04.72 OR positive C. difficile lab test AND CDI-specific antibiotic (metronidazole, oral vancomycin, fidaxomicin)* | Sensitivity | 100 (94 to 100) |
|  | Specificity | 89 (81 to 95) |
|  | PPV | 88 (78 to 94) |
|  | NPV | 100 (95 to 100) |
|  | Accuracy | 94 (88 to 97) |
| **<u>Code required:</u>**<br>*ICD-10-CM code A04.72 AND CDI-specific antibiotic* | Sensitivity | 93 (84 to 98) |
|  | Specificity | 94 (87 to 98) |
|  | PPV | 93 (84 to 98) |
|  | NPV | 94 (87 to 98) |
|  | Accuracy | 94 (88 to 97) |
| **<u>Positive test required:</u>**<br>*Positive C. difficile lab test AND CDI-specific antibiotic* | Sensitivity | 100 (94 to 100) |
|  | Specificity | 94 (87 to 98) |
|  | PPV | 93 (85 to 98) |
|  | NPV | 100 (96 to 100) |
|  | Accuracy | 97 (93 to 99) |
| **<u>Any CDI drug only:</u>**<br>*CDI-specific antibiotic (metronidazole, oral vancomycin, fidaxomicin), other criteria optional* | Sensitivity | 74 (64 to 83) |
|  | Specificity | 73 (63 to 82) |
|  | PPV | 74 (64 to 83) |
|  | NPV | 100 (95 to 100) |
|  | Accuracy | 85 (78 to 90) |
| **<u>Oral vancomycin only:</u>** *other criteria optional* | Sensitivity | 100 (94 to 100) |
|  | Specificity | 79 (69 to 87) |
|  | PPV | 79 (68 to 87) |
|  | NPV | 100 (94 to 100) |
|  | Accuracy | 88 (82 to 93) |
| **<u>Positive C. difficile test only:</u>** *other criteria optional* | Sensitivity | 100 (94 to 100) |
|  | Specificity | 83 (74 to 90) |
|  | PPV | 82 (72 to 90) |
|  | NPV | 100 (95 to 100) |
|  | Accuracy | 91 (85 to 95) |

Abbreviations: CI, confidence interval; PPV, positive predictive value; NPV, negative predictive value

## Discussion

Our study demonstrates that we can accurately identify inpatient HA-CDI cases using a combination of drug administration, laboratory testing, and ICD-10 code data from the EHR. Our algorithm was especially adept at capturing true positives (100% sensitivity). Additionally, 100% of those classified as CDI negative by our algorithm were truly negative (NPV). Requiring a positive *C. difficile* laboratory test improved the diagnostic accuracy even further by avoiding 5 false positives, and the combination of the three algorithm components performed better than any individual component or pair of components.

Our study contributes to the limited body of literature around construction of multi-component CDI case definitions, as much of the existing literature examines the utility of using a single component to detect cases. For example, a study my Litvin et al. reports on a “pseudo-outbreak” of CDI due to a faulty assay lot leading to a 32% perceived increase in CDI incidence at a facility (23). This illustrates the importance of multi-component algorithms. Planche and Wilcox highlight the varying sensitivity and specificity of different *C. difficile* laboratory tests alone (24). A multi-center study by Dubberke et al. reports that ICD-9-CM codes are an acceptable proxy for overall CDI burden, but fail to accurately capture HA-CDI burden (25). The authors note that the transition to ICD-10-CM codes was designed to more accurately delineate recurrent from non-recurrent, which is in agreement with an article by Deshpande et al. (26). A 2020 study in Canada by Pfister et al. reports that the ICD-10-CM code for non-recurrent CDI has a sensitivity of 85% and a positive predictive value of 80% while applied to a province-wide discharge database (27). Differences in coding practices could explain the heterogeneity in diagnostic performance across settings. Each of these studies identifies an important pitfall of single component case detection, thus motivating our validation study.

CDI-specific antibiotic treatment is another candidate for retrospective CDI case identification. While historically, metronidazole was the preferred treatment for non-severe CDI, oral vancomycin (and/or the more expensive fidaxomicin) is now the recommended treatment in many situations (13, 14). Though oral vancomycin is used exclusively for CDI, prophylaxis is becoming more common, so using oral vancomycin administration as a proxy for CDI can also overestimate the number of cases (15). We saw a lower specificity, positive predictive value, and overall percent accuracy when examining oral vancomycin in our data.

Our study is not without limitations. Because of CDI’s rarity in our patient population, it is highly unlikely that we would randomly find a “missing” CDI case among the sampled non-CDI group. While we calculated our power and sample size *a priori*, it is still possible that we are underestimating our denominator for our specificity calculations. However, this would not affect the sensitivity or positive predictive value of our algorithm. Additionally, our hospital has a low burden of HA-CDI compared to the national average (4.4 vs 8.3 case per 10,000 patient days according to a 2020 meta-analysis by Marra et al. (28)), so we might not be able to generalize our findings to setting with higher CDI burdens. Finally, while we leveraged our group’s expertise to establish our gold standard, the possibility of human error due to the clinical complexity of CDI remains. A major strength of our study is data availability. Our pharmacy research repository has comprehensive, longitudinal data that is easily linked to each patient, and has been validated in prior epidemiological studies (16).

Our study has important implications. Our CDI case definition algorithm can be applied as a gold standard to readily available EHR information to accurately detect CDI cases. This improves on commonly used methods like CDI LabID events. Accurate identification of CDI cases is crucial in the patient care setting (i.e., only prescribing treatment for true CDI patients), in research settings (i.e., quick and accurate CDI classification for retrospective studies), and for CDI tracking (reporting accurate facility-wide CDI incidence). Our algorithm was able to detect CDI cases with 100 percent sensitivity and high overall accuracy. Requiring a positivity laboratory test improved our algorithm’s diagnostic accuracy even further. Therefore, we recommend consideration of both a CDI-specific medication and a positive laboratory test (hospital day 4 or later) as the new gold standard when classifying HA-CDI cases from EHR data.

## Data Availability

All data produced in the present study are available upon reasonable request to the authors and completion of a data use agreement.

## Funding

This project received support from NIH grant UL1TR002369 and RL5GM118963

## Notes

### Competing Interest Statement

The authors have declared no competing interest.

### Author Declarations

The Institutional Review Board of Oregon Health & Science University gave ethical approval for this work

